# Development of a Mobile Application to Represent Food Intake in Inpatients: Dietary Data Systematization

**DOI:** 10.1101/2022.10.17.22278856

**Authors:** ARJO Molter, PE Brazil, CF Fonseca, AC Bacelo

## Abstract

**Background:** Nutritional risk situations related to decreased food intake in the hospital environment hinder nutritional care and increase malnutrition in hospitalized patients and are often associated with increased morbidity and mortality.

**Objective:** To develop and test the reliability of a mobile application as a virtual instrument to assess the acceptability and quality of hospital diets for inpatients.

**Methods:** This intra- and interobserver development and reliability study investigated an in-hospital food intake monitoring application based on a validated instrument for patients with infectious diseases who were treated at the Evandro Chagas National Institute of Infectious Diseases (INI/FIOCRUZ). The instrument was sequentially administered to patients 48 h after admission to INI hospital units using the printed instrument (paper) and the tested digital application (ARIETI) simultaneously. The tested reliability factor was the consistency of the method in the digital platform, checking whether the application provided equivalent data to the paper instrument, and finally a statistical analysis plan was performed in the R platform version 4.2.0. This project was authorized by the FIOCRUZ/INI Research Ethics Committee under CAAE 35379820.4.0000.5262.

**Results:** The ARIETI was developed and tested for reliability in 70 participants, showing similar ability to show caloric intake in Kcal (p = 1.72 E-03), protein intake (g) (p = 0.006362), the proportion of caloric intake (p = 1.54 E-05), and the protein intake (p = 0.003501) relative to the prescribed goal. The application was superior to the paper-based instrument, accelerating the diagnosis of nutritional risk based on food intake by up to 250 s (50–350 min).

**Conclusions:** The ARIETI optimized the time between diagnosis of nutritional risk related to dietary intake and the nutritionist’s decision making, showing an improved ability to maintain information quality compared to the paper-based instrument.

## Introduction

Malnutrition is an important public health problem worldwide. The risk of decreased food intake in the hospital environment hinders nutritional care, representing an important malnutrition factor (1), increase the morbidity rate, and extend the hospital stay (2); this, food intake is crucial for analyzing the patient’s nutritional, clinical, and hospital conditions.

Factors such as the underlying disease, coexisting acute or chronic comorbidities, medication side effects, physical inactivity, food supply deficiency and/or intake, psychological factors, inappetence or loss of the sense of taste, inability to ingest food or malabsorption, and the hospital environment itself, as well as frequent negligence of health teams regarding nutritional aspects to the detriment of other car can influence the nutritional status before, during, and even after hospitalization (3,4).

Proper monitoring of dietary acceptance ensures correct nutrient intake to hospitalized patients and can preserve or recover their nutritional status (5). Patients with infectious diseases such as AIDS, tuberculosis, Chagas and paracoccidioidomycosis, and coronavirus disease (COVID), among others, have increased nutritional demand increased and impaired food intake capacity as a results of inflammatory cascades (6,7).

Few validated instruments for monitoring in-hospital food intake can be quickly administered. However, Silva (8) developed a consistent instrument with a good correlation with the weighing method, the gold standard for dietary anamnesis, which requires no training in nutrition, only being literate(8). Despite these advantages, as this instrument is structured in paper, it carries the risk of document loss and transcription and/or analysis error regarding prescription(9). Moreover, using this instrument, the nutritionist has to manually calculate both the caloric and protein intake and adapt it to the prescription and patients’ nutritional needs to enable technical interpretation, risk diagnosis, and prescribed conduct adjustments.

Thus, the development of a digital solution based on Silva’s tool (8) simplifies and accelerates the diagnosis of in-hospital dietary intake by automatically calculating the food intake and the degree of adequacy or inadequacy according to the patient’s nutritional needs without the need for additional calculations and with greater information security.

Here we describe the process of improving the analog instrument (on paper) to develop an interactive and virtual data application for mobile devices and validate its use for in-hospital nutritional clinical practice.

## Methods Development

A digital solution for the diagnosis of nutritional risk based on in-hospital dietary intake was developed using multiplatform programming software, including Android and later IOS, in combination with C# and JSON technologies due to the high representativeness of the Android mobile platform in the Brazilian and global consumer market(10).

The digital solution developed was termed “Mobile Application to Represent the Food Intake of Inpatients” (ARIETI; Aplicativo móvel de Representação da Ingestão alimEntar de pacienTes Internados). The ARIETI is an improvement of the analog instrument (on paper) proposed by Silva(8), while still maintaining the humanized design and practical usability of the original instrument (Figure 1).

**Figure 1.**
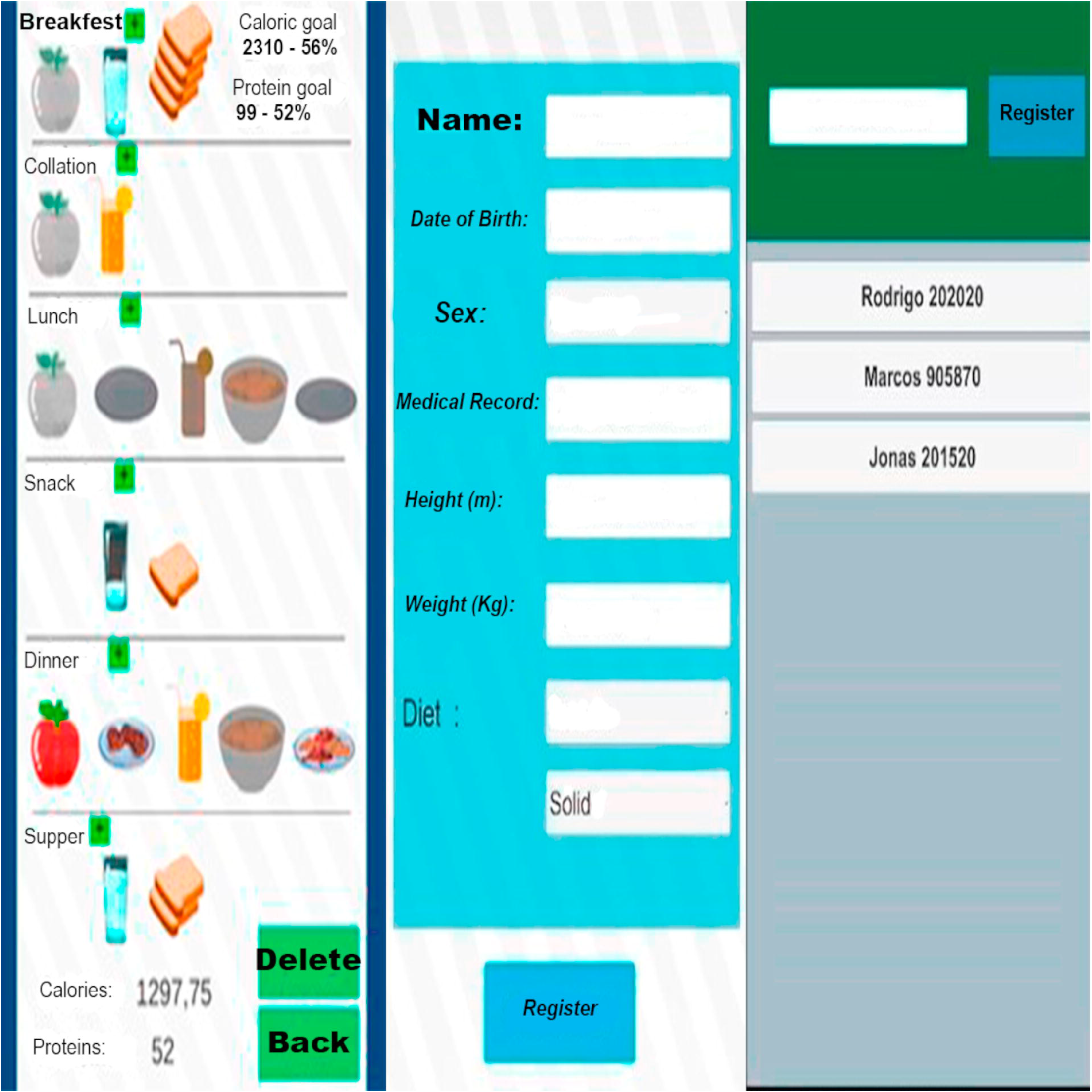
Application registration and data collection screens

The ARIETI was developed to identify variations in food intake calories, proteins, carbohydrates, and lipids, and to consider diet consistency variations, the dietary restrictions planned in the patient’s menu (sucrose, protein, or fats), and the partial or full acceptance of the different components of large and small meals (fruit, milk, bread, rice, beans, etc.), correlating them with the established therapeutic proposal (prescribed diet) and the patient’s nutritional goal (caloric and protein nutritional needs). The aim of the ARIETI was to help the nutritionist to identify nutritional risks related to adequate food intake inability, and to highlight the percentage of calorie or protein inadequacy and the reasons reported for the inability of partial or global intake to identify the need for dietary prescription adjustments. The ARIETI also optimizes the physical space for information storage, provides data security, and streamlines communication to minimize the prevalence of hospital malnutrition.

The ARIETI is an application (health app) that was developed using the Unity Software (Unity version 2019.1.1f1 (64-bit), 2019), which promotes more reliable data collection by making it simple and intuitive and can be easily used by any professional. ARIETI reduces the possibility of human calculation errors by aggregating all tools and algorithms, which optimizes the diagnosis of nutritional risk related to insufficient food intake, which is essential for more assertive decision making by the nutritionist.

### Embedded text

|  |  |  |
| --- | --- | --- |
| Name and last name | Name: Full name | Breakfast |
| Register | Date of birth: day/month/year | Caloric goal |
| Rodrigo | Sex: Male | Protein goal |
| Marcos | Medical record No.: Medical record No. | Collation |
| Jonas | Height (m): | Lunch |
|  | Weight (kg): | Snack |
|  | Diet: Light | Dinner |
|  | Solid | Supper |
|  | Register | Calories: |
|  |  | Proteins: |
|  |  | Delete |
|  |  | Back: |

### Study design and location

This observational, prospective study conducted a development and reliability assessment of a technological innovation tool for in-hospital dietary anamnesis, and was approved by the FIOCRUZ/INI Research Ethics Committee (CAAE No. 35379820.4.0000.5262). The tool was tested in patients hospitalized at the FIOCRUZ/INI.

#### Inclusion and exclusion criteria

The inclusion criteria were inpatients diagnosed with infectious disease who were awake, oriented and able to answer the instrument coherently, aged between 19 and 65 years, of both sexes, and who agreed to participate in the study.

The exclusion criteria were inpatients who were comatose or disoriented, with any diagnosed psychiatric disorder that could affect the coherence of the responses to the instrument, or with confirmed infectious disease in the first 24 h of hospitalization.

### Data collection

The food intake data of the participants were simultaneously collected in the printed form (Silva’s analog instrument, 2017) and in the ARIETI by two research collaborators who had experience in dietary assessment.

The time of bedside dietary assessment and the time between the inclusion of the participants’ responses and the diagnosis of nutritional risk secondary to the dietary intake pattern in the previous 24 h, both reported on paper and in the ARIETI, were entered into the RedCap platform. This was implemented to reduce data quality issues, allow dynamic data management (11), to help to ensure control and confidentiality, and to provide efficient data transfer into statistical analysis software such as the R-Project®.

The data collected by the ARIETI were stored on a MySQL server with and Hyper Text Transfer Protocol Secure (HTTPS) data encryption protocol to ensure protection; this protocol was chosen to protect the integrity and confidentiality of the data stored between the user’s hardware and the online application. All stored data are protected by the Transport Layer Security (TLS) protocol, which adds three additional layers of protection, including encryption, impossibility of corrupting data during transfer, and authentication, which ensures direct communication with the correct online application (12).

Recording food intake in the ARIETI allows a backup using an internet connection through the MySQL Workbench or using a USB platform between a computer, notebook, or tablet, thus enabling external storage and the creation of a functional relational database.

The sample size for the reliability test was 70 participants, considering a power of 0.8, and significance was considered if the p-value was ≤ 0.05. A flowchart of the study procedures is shown in Figure 2.

**Figure 2.**
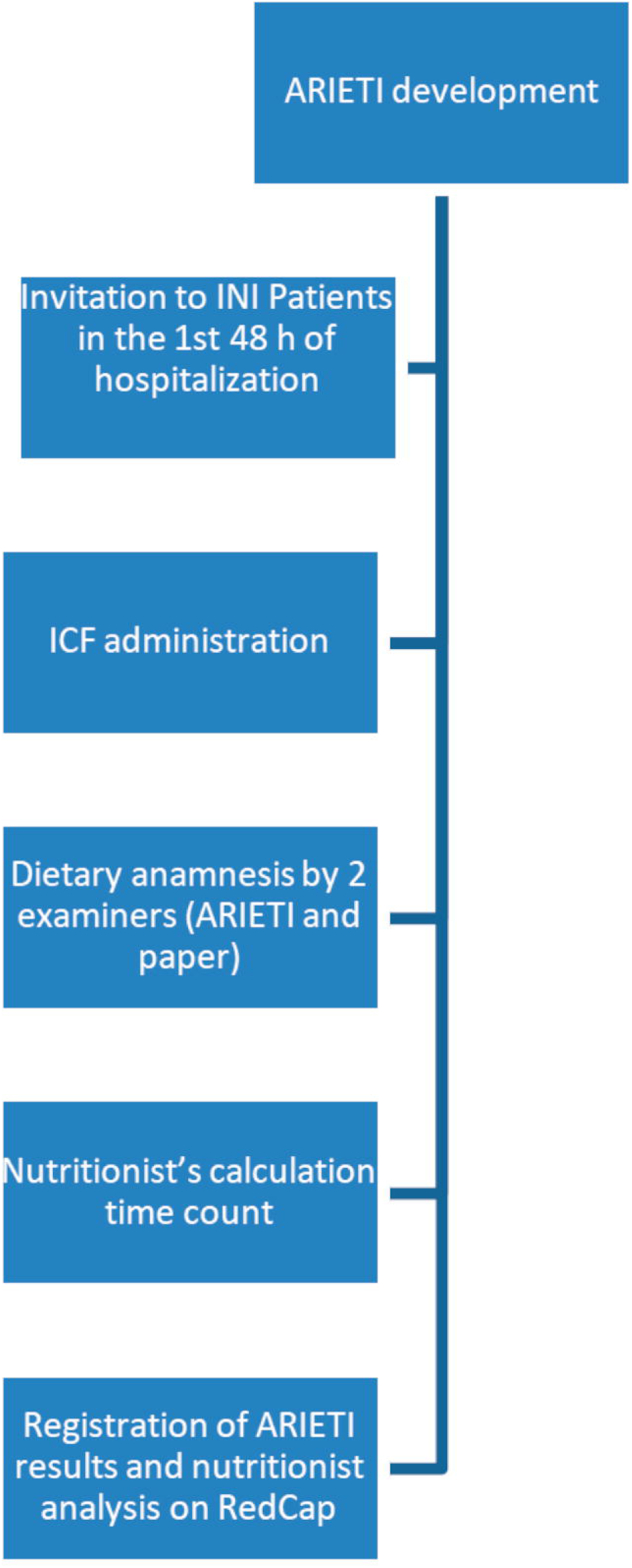
Flowchart of the study procedures

Embedded text: ARIETI development/Invitation to INI patients in the first 48 h of hospitalization/ICF administration/Dietary anamnesis by two examiners (ARIETI and paper)/Nutritionist’s calculation time count/Registration of ARIETI results and nutritionist analysis on RedCap. ARIETI: Aplicativo móvel de Representação da Ingestão alimEntar de pacienTes Internados; INI: Evandro Chagas National Institute of Infectious Diseases; ICF: informed consent form.

The reliability was verified by comparing the results of the dietary anamnesis between the paper and the ARIETI instruments. Therefore, in addition to data on food intake and adequacy results calculated by the algorithm, the ARIETI organizes information including name, age, sex, weight, height, calorie requirements (Kcal/kg of weight), and protein (g/kg of weight), according to the clinical decision of the prescribing nutritionist.

The data compared between the instruments included dietary data, anamnesis time (filling out the paper instrument and the ARIETI), and the time between the start of the bedside assessment and the clinical decision making.

### Embedded text

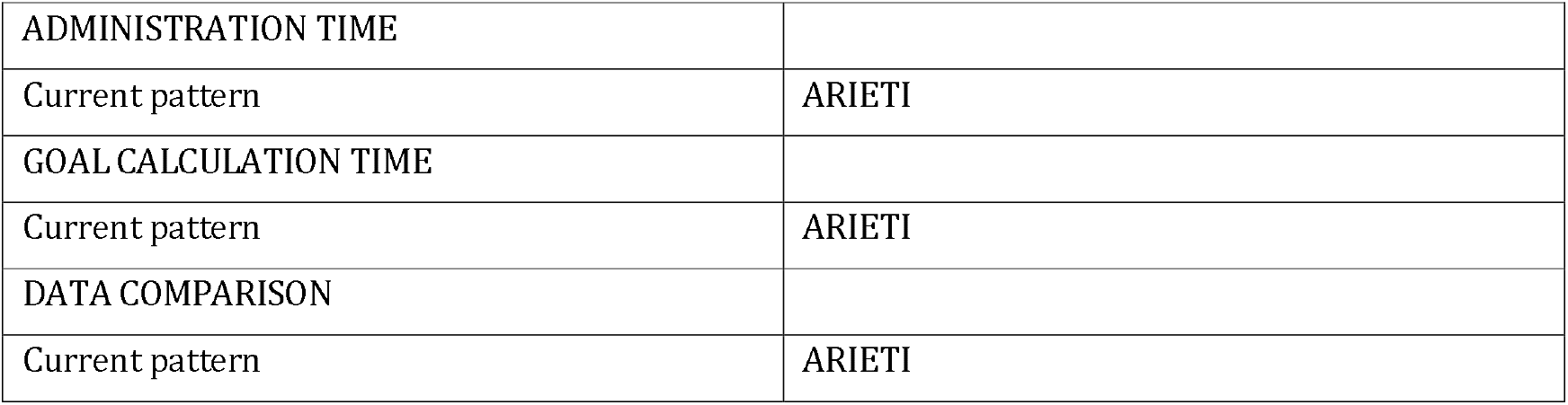

|  |  |
| --- | --- |
| ADMINISTRATION TIME |  |
| Current pattern | ARIETI |
| GOAL CALCULATION TIME |  |
| Current pattern | ARIETI |
| DATA COMPARISON |  |
| Current pattern | ARIETI |

The data were grouped by the following continuous quantitative variables, which were analyzed in both the paper and ARIETI:

⍰ Total Kcal: Total number of kilocalories ingested by the patient in 24 h of hospitalization.
⍰ Total Ptn: Total amount of protein ingested by the patient over 24 h of hospitalization.
⍰ Total Kcal goal: Percentage of the nutritional goal of the total number of kilocalories to be consumed by the patient, as stipulated by the nutritionist according to the patient’s nutritional status.
⍰ Total Ptn goal: Percentage of the nutritional goal of the total amount of protein to be consumed by the patient, as stipulated by the nutritionist according to the patient’s nutritional status.
⍰ Time diff paper ARIETI: Difference between the administration time and decision making for the two instruments.

### Statistical analysis plan

The data distribution was assessed using histograms and the Shapiro–Wilk test. The analysis of normality and the statistical curve trend guided the selection of hypothesis test (Pearson’s R or Wilcoxon’s Test) to evaluate the agreement between the printed instrument and the ARIETI (health app).

Data modeling was performed to compare the filling-in time and decision making. The Bland–Altman plot was used for visual comparison of the data.

All data will be analyzed using the R-Project® software version 4.2.0 and differences was considered at a power of 0.8 and significant if the p-value was ≤ 0.05.

## Results

The ARIETI optimized the time between the diagnosis of nutritional risk related to dietary intake and the nutritionist’s decision making, improving the quality of information compared to the paper-based instrument.

The calorie and protein intake recorded on paper and in the ARIETI were similar, as shown in the histogram with trend curves to the right in both instruments, for both calorie and protein intake (Figure 3), and in the boxplot, which shows a slightly smaller amplitude and median total calorie in the ARIETI than in the paper version (Figure 4).

**Figure 3.**
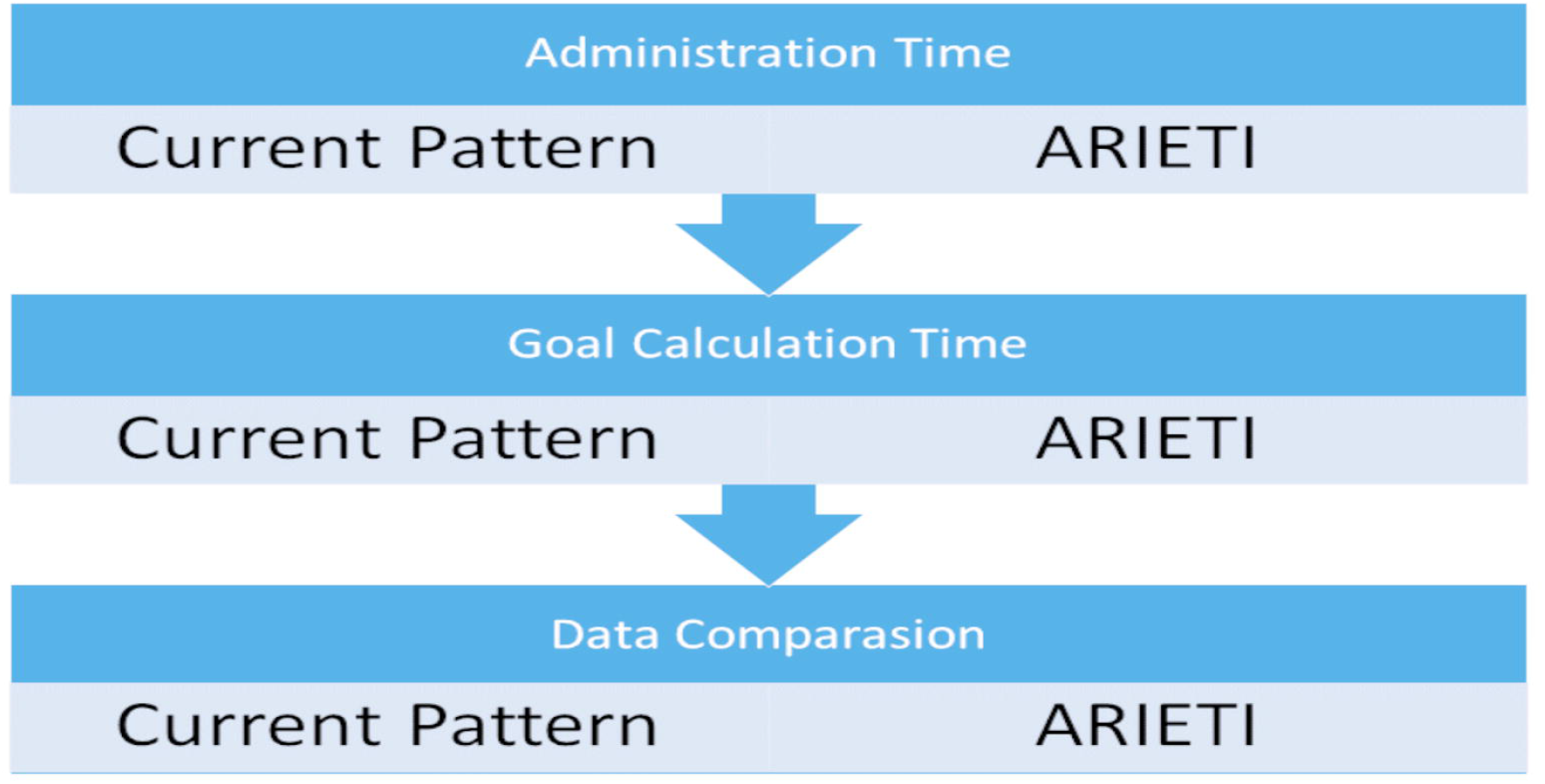
Methodological path for reliability analysis

**Figure 4.**
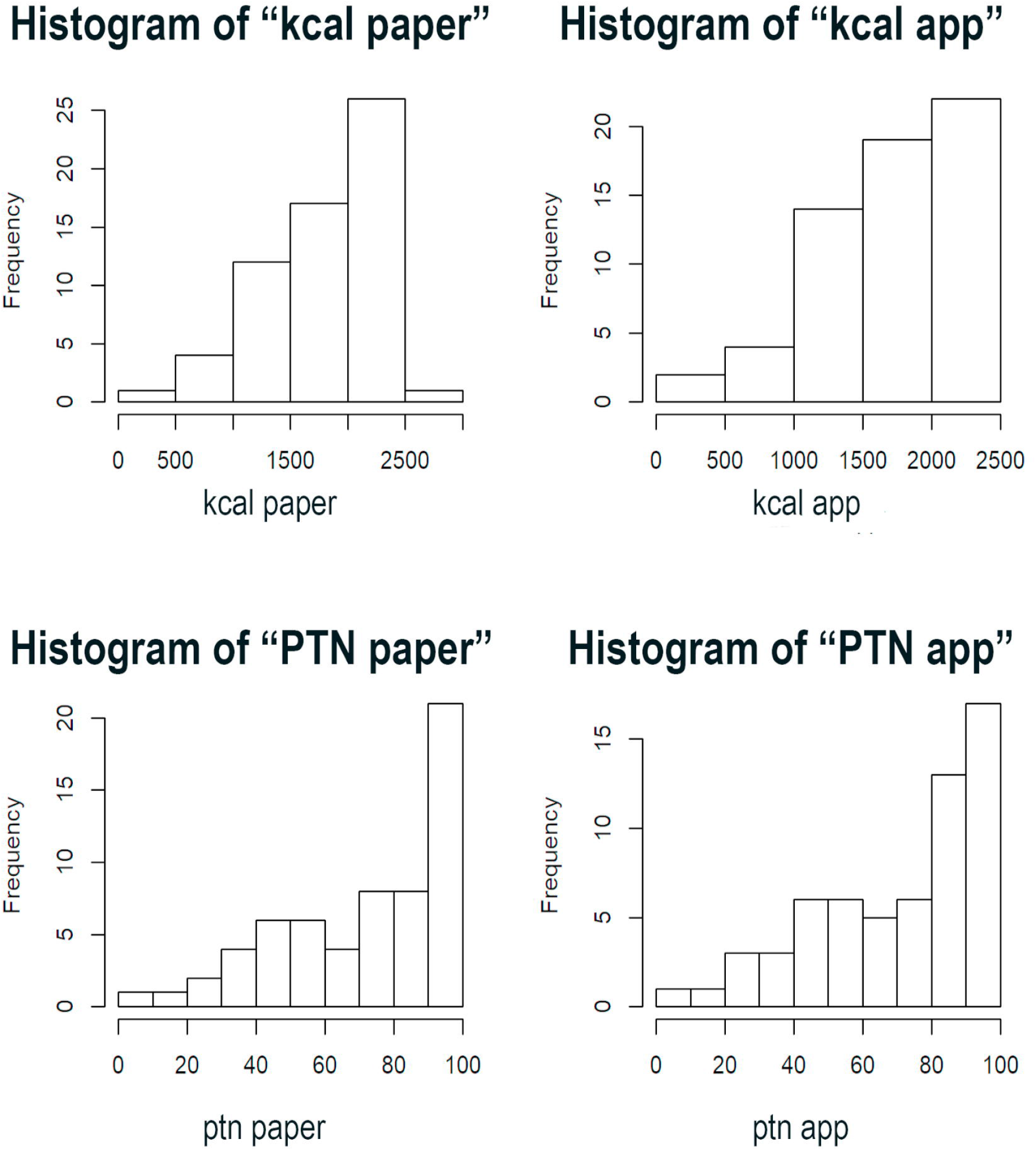
Histograms showing the comparison of calorie and protein intake between the paper instrument and the ARIETI

The outliers observed in the proportion of food intake capacity regarding calorie goals calculated by the ARIETI were the same as those calculated manually by the nutritionists.

There was no relevant statistical difference between the two instruments (paper and ARIETI) regarding the dietary anamnesis variables investigated for the diagnosis of nutritional risk associated with food intake, proving the null hypothesis (H0), thereby showing no significant difference between the diagnostic result established by the paper instrument and the ARIETI.

The p-values were < 0.05 (Table 2) and relevant considering the values obtained from the analysis of pairs of the same variable between the two instruments regarding food intake of kilocalories, proteins, and their respective nutritional goals of 1.72E-03, 0.006362, 1.54E-05, and 0.003501 for the Wilcoxon’s test and < 2.2e-16 for Pearson’s correlation. These results prove the statistically similar performance of the application compared to the paper instrument in all variables analyzed.

**Table 1.**
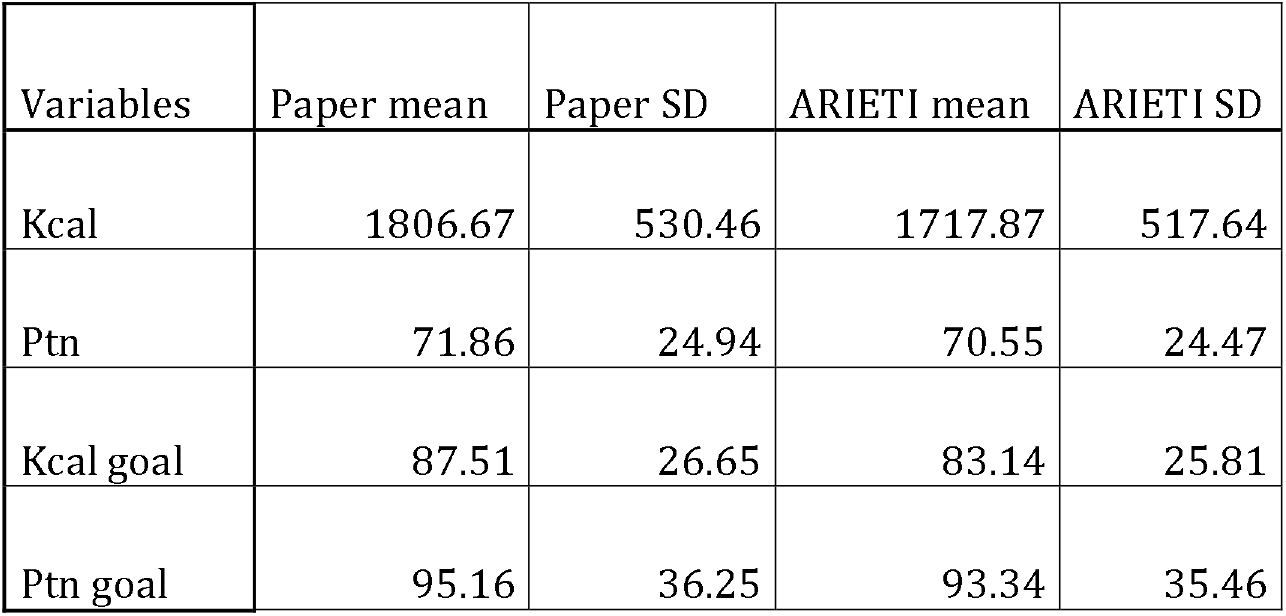
Means and standard deviation of protein intake (g), number of calories, and percentage of nutritional goals achieved calculated by the two instruments.

| Variables | Paper mean | Paper SD | ARIETI mean | ARIETI SD |
| --- | --- | --- | --- | --- |
| Kcal | 1806.67 | 530.46 | 1717.87 | 517.64 |
| Ptn | 71.86 | 24.94 | 70.55 | 24.47 |
| Kcal goal | 87.51 | 26.65 | 83.14 | 25.81 |
| Ptn goal | 95.16 | 36.25 | 93.34 | 35.46 |

**Table 2.**
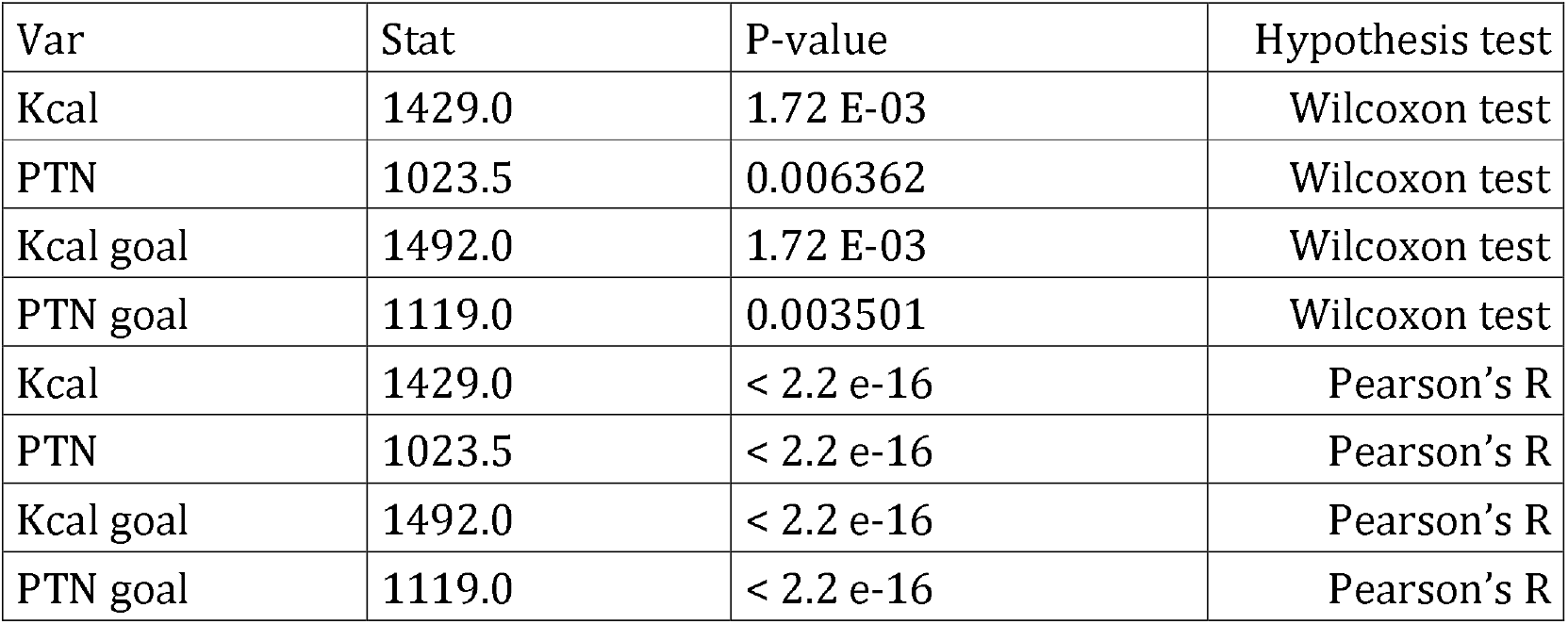
Ability of the ARIETI to report the same caloric and protein intake values and to propose the same diagnosis of nutritional risk associated with caloric and protein intake capacity as the paper-based instrument, according to the appropriate hypothesis tests. Stat: Hypothesis test calculated for the same variable in both instruments (app and paper).

| Var | Stat | P-value | Hypothesis test |
| --- | --- | --- | --- |
| Kcal | 1429.0 | 1.72 E-03 | Wilcoxon test |
| PTN | 1023.5 | 0.006362 | Wilcoxon test |
| Kcal goal | 1492.0 | 1.72 E-03 | Wilcoxon test |
| PTN goal | 1119.0 | 0.003501 | Wilcoxon test |
| Kcal | 1429.0 | < 2.2 e-16 | Pearson's R |
| PTN | 1023.5 | < 2.2 e-16 | Pearson's R |
| Kcal goal | 1492.0 | < 2.2 e-16 | Pearson's R |
| PTN goal | 1119.0 | < 2.2 e-16 | Pearson's R |

The Bland–Altman plot demonstrates the calculated mean of the two instruments and the measurement differences between them. The black line represents the mean nutritional goals and caloric and protein intake differences between the two instruments, while the two red dotted lines represent the 95% confidence interval limits for the mean difference.

Embedded text: META = TARGET/PAPEL = PAPER/E = AND

This graphic visualization method shows that data dispersion follows the mean differences between the variables studied with the two instruments used, following the expectancy for the trend line in the comparison between them. The dispersion showed a low number of outliers, with the data respecting their statistical equivalence, i.e., the instruments represent food intake with the same quality and capacity.

The ARIETI showed good reliability for dietary anamnesis, as can be confirmed in the linear dispersion presented in Figure 6A–D (Bland–Altman plots on the distribution and mean nutritional goal, and calorie and protein intake goals, according to the paper instrument and ARIETI).

**Figure 5.**
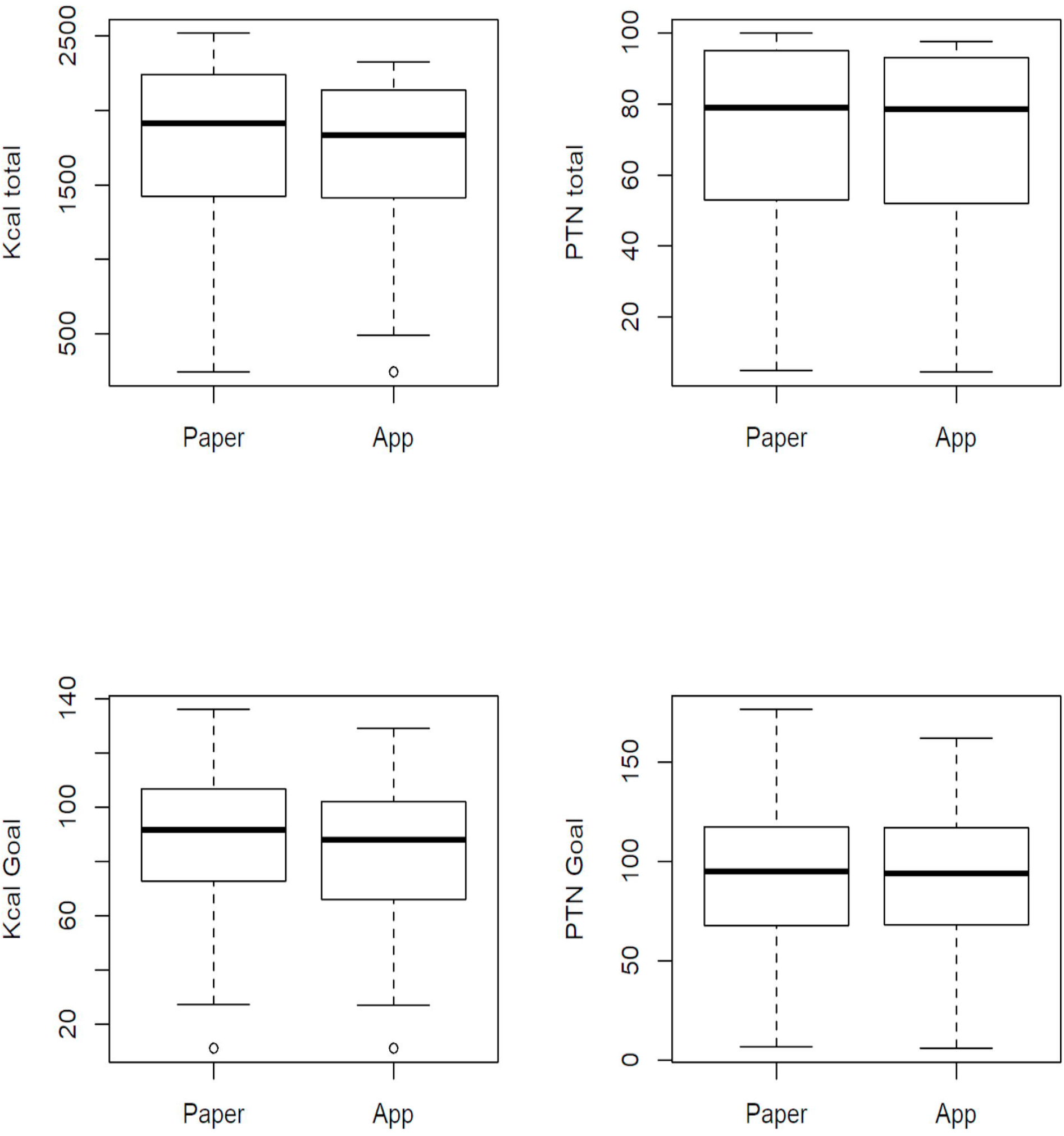
Boxplots showing comparisons between variables

**Figure 6.**
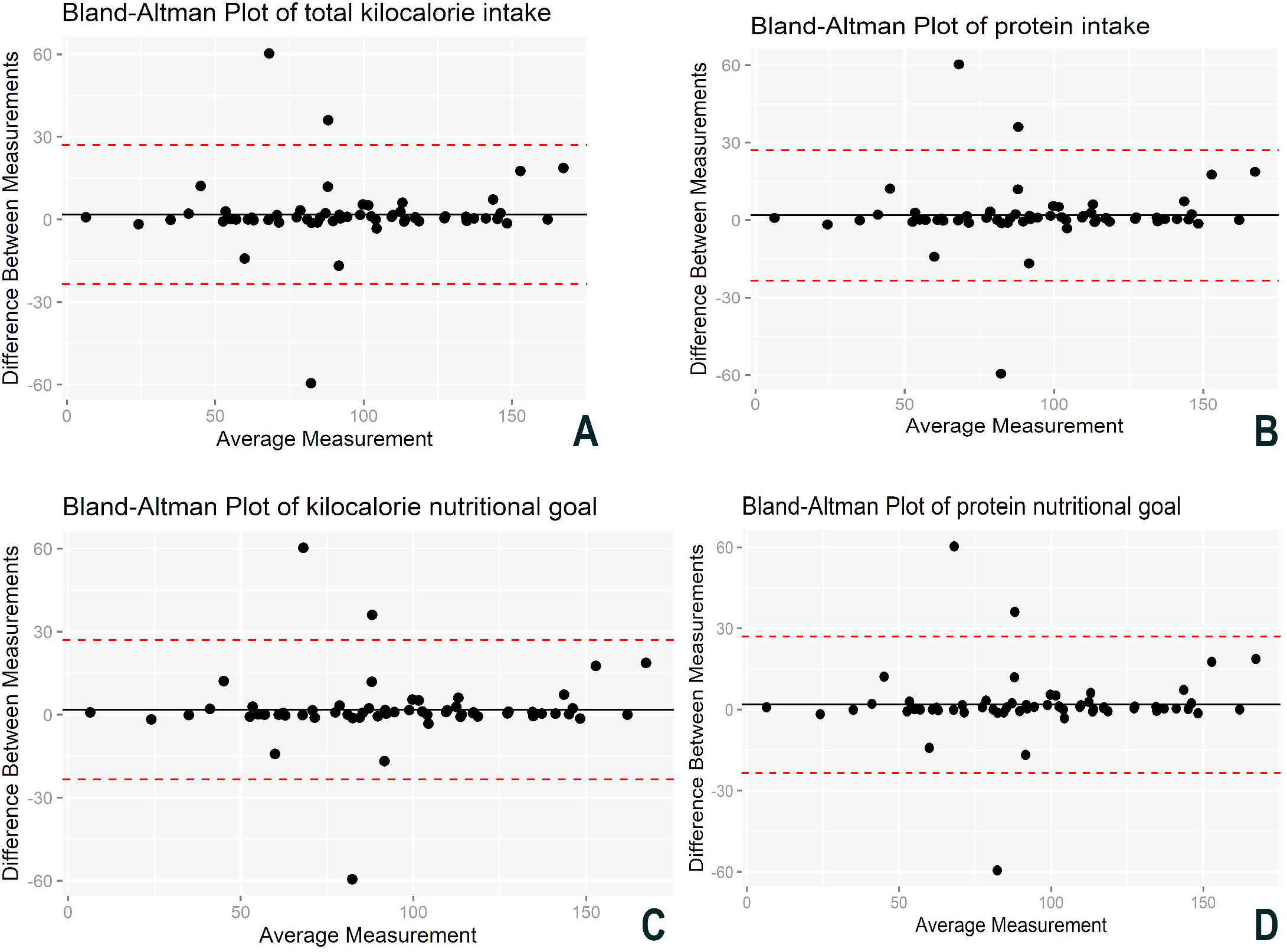
Bland–Altman Plot of total kilocalorie intake (A), total protein intake (B), kilocalorie nutritional goal (C), and protein nutritional goal (D) measured by the paper instrument and the virtual application. Embedded text: META = TARGET/PAPEL = PAPER/E = AND

As for the different times required between administration and decision making by the nutrition professional according to the patient’s nutritional diagnosis, the ARIETI accelerated the process by approximately 150 s (Figure 7).

**Figure 7.**
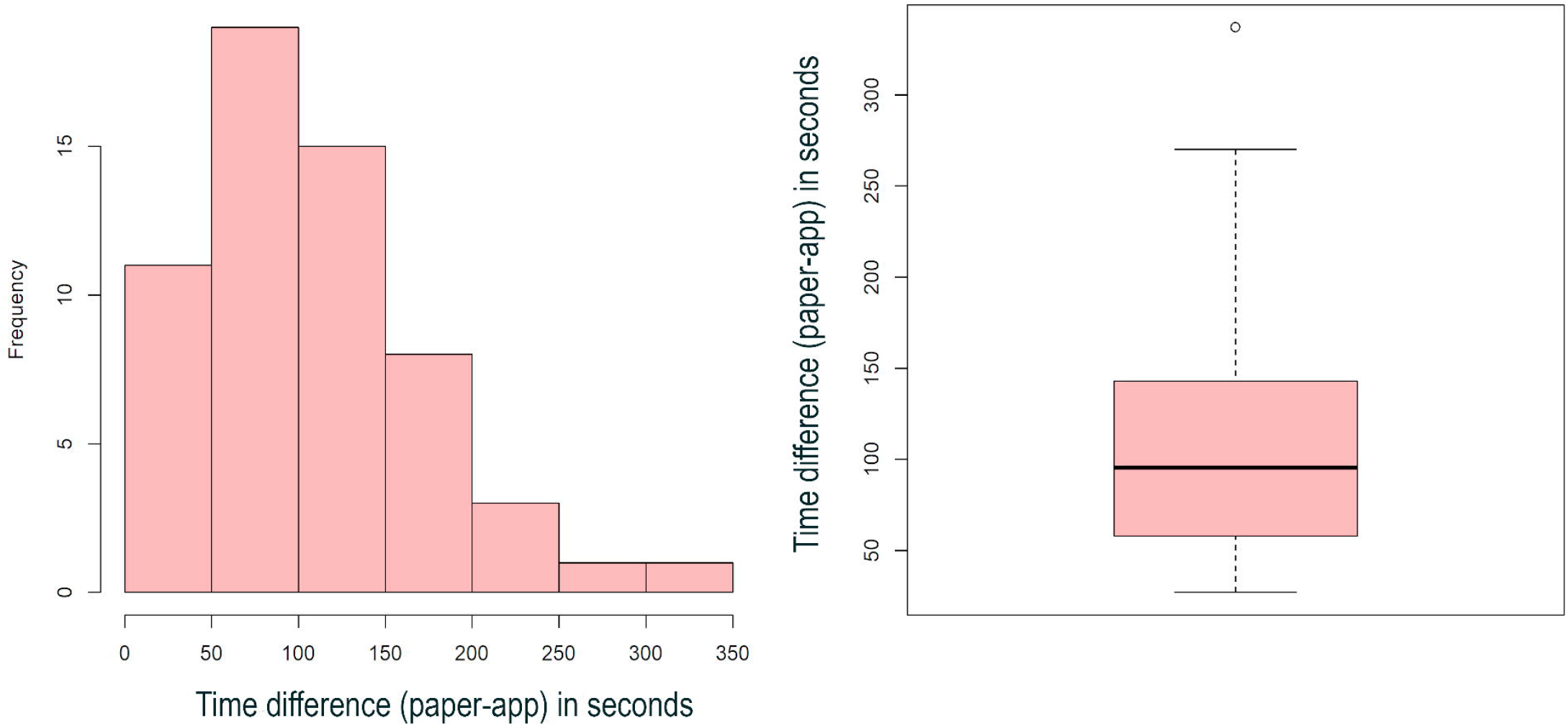
Analysis of the difference between decision-making times Embedded text: Time difference (paper-app) in seconds

In some situations, the time used for calculations and decision making related to the nutritional diagnosis of each patient (Figure 7) using the paper instrument was approximately ± 250 s greater when performed manually.

Embedded text: Time difference (paper-app) in seconds

## Discussion

In this study, we aimed to demonstrate the development phases and reliability of the smartphone application ARIETI, a digital solution for in-hospital clinical nutritional practice.

The reliability analysis of the developed digital solution showed the following main findings: a) the ARIETI accelerated the time to identify nutritional risk associated with food intake compared to the paper-based instrument; b) the ARIETI was reliable and could replace the paper-based dietary anamnesis instrument that is currently validated for hospital settings.

The interactive and humanized model (Figure 1) of the proposed instrument (ARIETI) optimizes dietary intake diagnostic activities, saves professional time, and avoids rework. Additionally, this form of management contributes to appropriate and agile adjustment of hospital diets and better monitoring of clinical nutritional progression.

Food intake assessment is crucial to the dietary management of inpatients(13). Monitoring and recording the entire dietary content ingested by the patient are important components of their nutritional care, especially for those considered to be at higher nutritional risk (14,15).

The gold standard for assessing the acceptance of hospital diet in clinical studies is the weighing of food remnants that are not ingested by patients, (16–20); however, this is often unfeasible in the routine assessment of inpatients as it requires a longer time and greater number of professionals. Therefore, the instrument validated by Silva (8) was compared with weighing as the basis for developing the ARIETI.

The time to administer the methods, which consists of completing the paper and virtual forms in the application, was equal because to the completion time depended directly on the patient’s response time at the bedside. However, the time to make the necessary calculations for the nutritional diagnosis of the patient showed a slight difference (Figure 6).

The difference between the time of calculation and the decision on the patient’s nutritional diagnosis is crucial considering that the virtual application provides the results of this analysis in real time at the bedside, fully streamlining this process, thereby having the potential to provide nutritional care actions with greater agility and in a shorter time interval (21).

In some cases, intercurrences, such as forgetting details of the anamnesis, may occur at the time of the clinical visit that influence the process of recording patient data before nutritional calculations. Given that the ARIETI returns the data that have already been calculated in real time, its long-term use can save a considerable amount of time spent on calculations, besides ensuring that human errors do not occur in the diagnostic operation of nutritional risk related to in-hospital food intake capacity.

The ARIETI was developed to modernize the method and make it significantly more practical and rapid to accelerate data collection, with the aim to translate into a continuous quality of care improvements to service users (22). Storing data digitally saves space and reduces costs compared to storage using paper and other materials, besides enabling access from anywhere in the world (23). Therefore, using an electronic system in the cloud, physical devices, or in backups for nutritional data collection improves data security, reliability, and accessibility.

The curve tends to the right (Figure 3), changing the normality of the distribution because the study included fewer patients with severe symptoms than with normal conditions(stabilized pathological condition). The severity of the condition affects the nutritional diagnosis given that patients with more severe disease tend to develop a nutritional deficit (24). Given this characteristic, we used the Wilcoxon test to analyze the relationship between the two methods.

The boxplot shows the data distribution and outliers, thereby providing a complementary means to develop a better perspective on the character of the dietary intake variables and nutritional goals. Additionally, the boxplot served as a comparative graphical layout of the two instruments. The discrete presence of outliers and the slight differences in the boxplot medians (Figure 4) may theorize as a cause a possible human calculation and measurement errors with the paper instrument (25).

As an alternative to the Student’s t-test, in which the samples follow a normal distribution, the Wilcoxon test proved the similarity of the medians of the calorie and protein food intake variables, in addition to the nutritional goals obtained. Thus, it is possible to assess the efficacy of the virtual method tested, ensuring the safe assessment of the nutritional diagnosis of inpatients through food intake analysis and established nutritional goals (26).

The power of statistical significance by Pearson’s correlation < 2.2e-16 (Table 2) for food intake and nutritional goal prove the linear trend of the positively correlated data given that the magnitudes remained similar according to the variations(27).

The Bland–Altman method demonstrated the dispersion between the individual means of food intake and nutritional goal variables, in addition to the individual differences between them, describing the agreement between the two methods (paper and ARIETI) considering quantitative variables. The assessment of the limits of agreement should include the clinical perspective, i.e., to determine whether the differences given by the limits can be considered acceptable (28).

The Bland–Altman method should be used in full when it is necessary to assess the agreement between two methods, including limits of agreement and their confidence intervals. If case a change becomes essential, the data must be corrected to obtain a reliable and cohesive conclusion (29). There was good dispersion of the data given that the results followed the trend lines of the means and most of the distribution was within the limits of the 95% confidence intervals (Figure 6A–D).

### Key findings

The ARIETI demonstrated high reliability and efficacy as a virtual instrument to diagnose nutritional risk by dietary analysis and can therefore be reliably implemented in clinical practice.

### Limitations

The ARIETI requires internet access given the required communication with the relational database (SQL), and has been developed for the Android operating system (OS) versions 4.1 or higher; devices with other OS versions cannot run the developed software, as well as devices with very outdated Android systems.

## Conclusion

The ARIETI increased productivity, allowing remote access, optimizing the appointment time, and eliminating the difficulties and limitations of a paper instrument, such as loss, and key element transcription or analysis errors in food intake control.

The use of technology focused on improving management, integrating information, and promoting efficiency gains allows health professionals to focus on their main objective, prescribing proper hospital diets and improving the efficiency of the therapeutic and nutritional processes in the hospital environment.

## Data Availability

Data cannot be shared publicly because of Informed Consent Form that guards the participants information and data. Data are available from the Instituto Nacional de Infectologia (contact via 55 (21) 3865-9602) for researchers who meet the criteria for access to confidential data. This observational, prospective study conducted a development and reliability assessment of a technological innovation tool for in-hospital dietary anamnesis, and was approved by the FIOCRUZ/INI Research Ethics Committee (CAAE No. 35379820.4.0000.5262). The tool was tested in patients hospitalized at the FIOCRUZ/INI.

## Acknowledgments

The authors wish to thank Rodrigo Silva de Oliveira, PhD, Programming Professor, for providing technical IT support.

## Authors’ contributions

BAC, BPE: Study conception and design; MARJO: Data collection; MARJO, BPE: Contributed to data or analysis tools; MARJO: Data analysis; MARJO, BAC: Manuscript writing; FCF: Code analysis and technical consulting.

## Abbreviations

App: Application
ARIETI: Mobile application to represent the food intake of inpatients
Diff: Difference
STD: Sexually transmitted diseases
H0: Null hypothesis
HIV: Human immunodeficiency virus
HTLV: Human cell lymphotropic virus
HTTPS: Hyper Text Transfer Protocol
INI: Infectious Diseases Institute
Kcal: Kilocalorie
Kcal goal: Nutritional total kilocalorie intake goal for the patient
Kcal total: Total kilocalorie ingested by the patient
OS: Operating system
PTN: Protein
PTN Goal: Nutritional total protein intake goal for the patient
PTN total: Total protein ingested by the patient
Stat: Test statistics
TLS: Transport Layer Security
Var: Variable

## Appendix

### Informed Consent Form

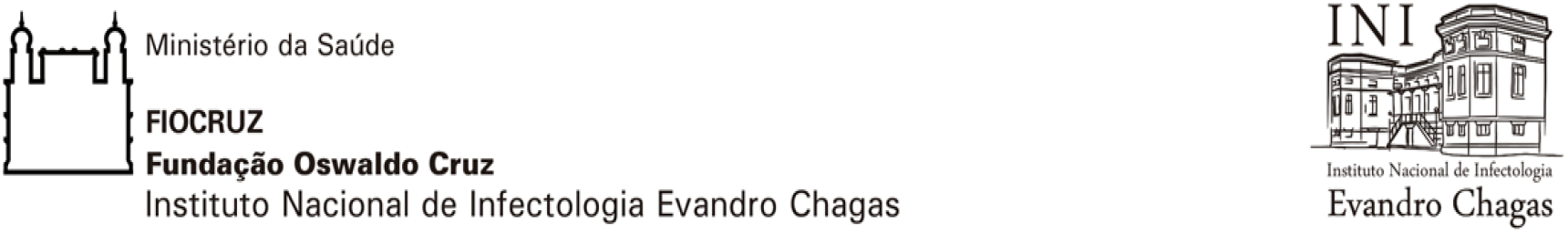

### INFORMED CONSENT FORM

#### Project: DEVELOPMENT OF A MOBILE APPLICATION TO REPRESENT FOOD INTAKE IN INPATIENTS: A RELIABILITY TEST

Head researchers: Alan Renier Jamal Occhioni Molter (INI/Fiocruz Teaching/Nutrition Service); Claudio Fico Fonseca (School of Engineering - UVA); Marlete Pereira da Silva and Patrícia Dias de Brito (INI/Fiocruz Nutrition Service), Pedro Emmanuel Brasil (LAB), and Adriana Costa Bacelo (INI/Fiocruz Nutrition Service) Telephone No.: (21) 3865-9602 (INI/Fiocruz Nutrition Service)

You are being invited to participate in this research, which aims to develop an application to be used as an instrument to assess the acceptance of hospital diets, i.e., to assess how much of the food that inpatients receive they are actually able to eat. You are being invited because you are hospitalized, awake, oriented, and able to feed yourself orally (by mouth), but your participation is voluntary and not mandatory. Please do not feel that you have to participate in this study, if you do not, you will still continue to receive adequate food and treatment.

#### Research objective

To verify the reliability of a digital instrument (a form structured through a digital application for use with a mobile phone or computer) to assess the acceptance of hospital diets compared to the paper instrument (printed form) that has already been validated.

#### Participation in the study

If you agree to participate in this study, you will answer the questions on this form to the nutritionist after the first 48 h of hospitalization. Two professionals will record your answers simultaneously; one will record your answers on paper, and the other will record your answers directly in the application installed in the mobile phone – this should last 10–15 min. If you are discharged and readmitted within the study period (July to December 2020), you may participate more than once.

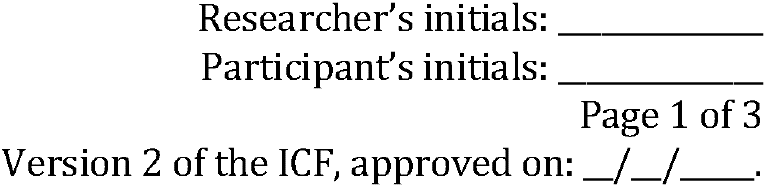

#### Risks and discomfort

Your risk in this research corresponds to the possibility of breach of confidentiality, so, to minimize this risk, your information will be recorded without personal identification, that is, the initials of your name, project inclusion number, and registration number in the institution; your full name and document number will never be recorded. The discomfort expected to be associated with this study is considered to related to the embarrassment of answering questions about your ability to accept the food received in the first hours of your hospital stay, especially in cases of readmission, because you will answer the same questions twice; however, you may stop participating at any time, following which, you will be immediately removed from the study and will not have to answer any more questions.

Please note that you have the right to claim compensation in the event of any damages resulting from this research, i.e., if you consider yourself to have been harmed in anyway as a result of your participation in this study.

#### Confidentiality

The researchers guarantee that your privacy will be respected, i.e., your name or any other identifiable data will be kept confidential.

#### Refusal guarantee

You may refuse to participate in the study or withdraw your consent at any time without having to justify it, and you will not undergo any food or assistance losses.

#### Benefit

You will not receive any personal benefit, but the outcome of the study may help improve to improve hospital food provision and assist with faster nutritional intervention for future patients.

#### Expenses and costs

You will have no expenses of any kind to participate in this research, nor will you receive anything for participating. You will participate only on 2 days for filling out the form and weighing meals while you are hospitalized, and you will not need to return to the institution another day.

#### Research results

At the end of the study, the researchers commit to disseminate the results obtained with this research in seminars, events, and scientific journals.

At any time, you may discuss any questions you may have regarding this study with the nutritionist, Adriana Costa Bacelo, at the following address: Serviço de Nutrição, INI/Fiocruz, Avenida Brasil 4365, Manguinhos, Rio de Janeiro (Tel.: 3865–9602).

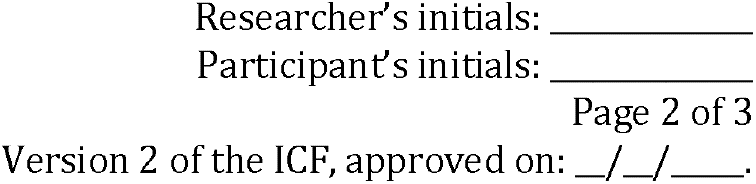

After agreeing to participate, you and the nutritionist will sign two copies of this form. One copy will remain with the research team and the other copy with you.

The procedures adopted in this research comply with the Human Ethics Research Criteria according to Resolution No. 466/2012 of the National Health Council and the study was approved by the INI Research Ethics Committee (REC).

The REC is a group that protects the interests of research participants and evaluates the conduct of research. You may also contact the REC to ask questions about your rights as a participant in this study at: INI/Fiocruz, Avenida Brasil 4365 - Manguinhos - Rio de Janeiro; Tel.: (21) 3865-9585.

Note: Do not sign this form if you still have questions relating to this study.

Informed Consent Form

I agree to participate in the research. I declare that I have received a copy of this consent form, and I authorize the research and the disclosure of the data obtained in this study.

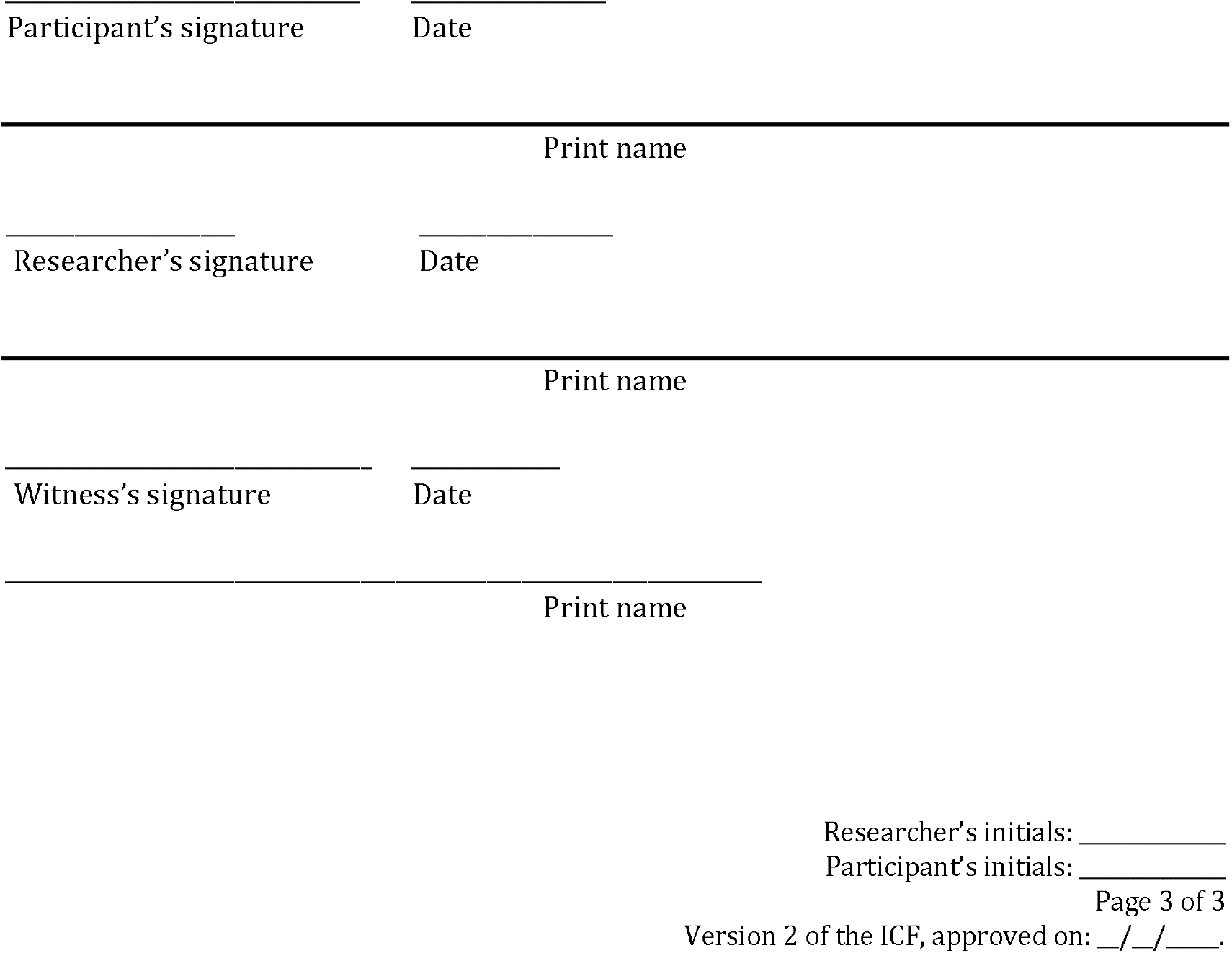

